# Trends, changes and determinants of medical attention received before death among deceased adults in India: Evidence from pooled cross-sectional survey data (2004-2018)

**DOI:** 10.1101/2022.05.22.22275424

**Authors:** Saddaf Naaz Akhtar, Nandita Saikia

**Author notes:** Address correspondence to Saddaf Naaz Akhtar. **Funding:** This study did not receive any funding. **Data availability:** The data is publicly available and can be accessed at https://www.mospi.gov.in/web/mospi/download-tables-data/-/reports/view/templateTwo/16202?q=TBDCAT.

## Abstract

**Background:** India is coping an ageing population without an adequate medical care service. However, trends, changes and factors of receiving medical attention before death remains unknown. Therefore, we intend to examine the trends, changes and determinants of medical attention received before death among the deceased adults in India.

**Methods:** Our study has used a nationally representative pooled-cross-sectional survey data from2004 to 2017-18. We performed univariate, bivariate and multivariate analyses. We have examined the trends from 2004 to 2017-18. Then we estimated the absolute changes in medical attention rate (MAR) received before death by the 45+ deceased adults for the time period between 2004-2005 & 2017-2018 and 2014 & 2017-2018. Lastly, we applied binary logistic regression analysis to identify the effect of significant predictor variables on the medical attention received before death.

**Results:** Our study has revealed that there has been marginal increase (0.74%) in the overall trend in medical attention received before death among 45+ adults from 2004 to 2017-18. Females, rural residence are showing negative change in receiving medical attention. However, our finding has showed that young-old, middle-old and oldest-old have significantly lower odds of receiving medical attention compared to middle-aged adults. Surprisingly, our result has revealed that Southern and Western regions are found to have significantly less likely to receive medical attention before death among deceased adults which is a striking finding.

**Conclusions:** Therefore, there is an urgent need to establish a primary health center at community level that play an important role in order to meet the comprehensive requirements of middle-aged and older adults in India. It would be helpful to develop and execute the health policies and programs that would enable India to achieve healthy aging in India at national and subnational levels, as it is a key element of public health challenges.

## Introduction

In India, research on medical attention among adult or old-age mortality is scanty since the majority of previous research focused on infant, child, and maternal mortality in the recent past(Bhat, 1987; Bora & Saikia, 2018; Saikia, Shkolnikov, et al., 2016). However, post-National Rural Health Mission (NRHM) program, India has been experiencing a substantial increase in life expectancy at birth due to reduction in fertility rate as well improvement reduction in the under-five mortality rate. Increasingly, old age morality and health care indicators are becoming indicators of social inequality and health and well-being in India. One such old age health care indicator is receiving medical attention at death. However, there is about 48.5% deaths occurred in 2019 who has received medical attention before death either in Govt. hospitals or private hospitals in India (SRS Report, 2019).

The term ‘medical attention’ received before death is defined as “a level of medical treatment or care where a doctor or physician delivers inpatient care or active treatment of chronic, obstetrical, surgical, medical, rehabilitative or mental illnesses that necessitate physician monitoring, diagnosis, and daily therapy or medication before death. (Law Insider, 2022). It is an important indicator that enables us to understand mortality causes related to medical care received could provide insight into the vital role of tackling one of the most pressing challenges in the developing world. It would also help strengthen India’s health system by delivering adequate healthcare services, especially for middle-aged and older adults. However, there is very limited research presently available on the subject of mortality who received medical attention in India (Ghosh & Sengupta, 2016; Gupta & Sankar, 2004) and other developing countries (Alam et al., 2010; Gbeasor-Komlanvi et al., 2020).

Medical attention is one of the primary sources of future health improvements. However, receiving medical care or attention before death is not a prime concern in developed countries because most people are covered with health insurance and they have improved access to health care facilities. Meanwhile the epidemiological transition and change in the age structure of India’s population is evidencing aging population, introducing higher spending on health care demand is of prime significance. Previous studies in India revealed that health care equity is highly impacted by a lack of comprehensive medical insurance plans, resulting in a rise in the number of options for accessing medical health care in private health institutions (Bose, 2005; Prinja et al., 2012). While the cost of medical health care in India suggests a lack of health insurance coverage that expose the older people, mainly those belonging to lower socio-economic groups, to significant financial distress (Sahoo et al., 2021).

However, previous studies showed that many middle-aged and older adults forgo medical care, regardless of the potential medical advancement and they avoid seeking medical help, even when they knew they should (Kannan & Veazie, 2014; Leyva et al., 2020). While other studies may appear counterintuitive that people ignore seeking medical attention that could save their lives or alleviate their health problems (Kannan & Veazie, 2014; Leyva et al., 2020; Spleen et al., 2014). Recent studies in India have showed that older adults have a lower health-related quality of life (Akhtar & Saikia, 2022; Boro et al., 2021; Kumar & Kumar, 2022; Kundu et al., 2022), and have a higher prevalence of multiple chronic illness (Chauhan et al., 2022; Khan et al., 2021). Because of their greater medical needs, older persons may be more exposed to the negative health implications of postponing medical care.

Unfortunately, India does not have enough age-appropriate medical institutes or health centers especially for older people such as geriatric facilities at community level. Therefore, country is experiencing lack of medical care services at community level that can deliver adequate health care for all ages separately. While a recent study has confirmed that health insurance coverage is a key determinant that could attribute to the better health utilization in India (Ali et al., 2022). Instead, the primary focus is based on the proportion of deaths happened in health institute or hospitals vs. those outside hospitals.

It has been realized that there are many deaths which have occurred without even receiving an adequate medical attention, especially for middle-age adults as well as older adults. Therefore, studying medical attention received before death is a very important and serious public health issues which also has implications for health professionals and policymakers. This can also highlight the problems of lower death registrations in India and also provide an appropriate picture of the mortality improvements in the country. Hence, there is a need of robust and coherent research to reduce these gaps in the knowledge and action based on evidence-based policy is required, in order to understand the trends, change and determinants of medical attention received by the deceased adults.

Therefore, the present study intends to examine the trends, changes and determinants of medical attention received before death by deceased adults in India using pooled cross-sectional survey data.

## Methods

### Data Source

The present study has used pooled sample data from 60^th^, 71^st^ and 75^th^ rounds of the National Sample Survey Organizations (NSSO), under the Ministry of Statistics and Program Implementation (MOSPI) which were conducted in India during the year 2004-2005, 2014 and 2017-2018 respectively. The unit level data of the deceased adults of social-consumption on health, schedule 25.0 for 60^th^, 71^st^ and 75^th^ rounds. The data is collected by the NSSO which is a nationally represented data, which has adopted a two-stage stratified probability sampling. There were 73868, 65932 and 113823 households that were survey in the 60^th^, 71^st^ and 75^th^ rounds of NSSO. Out of which 1717, 2395 and 2537 deceased adults’ cases in the household. Since, our study is only based on 45 years and above deceased adults therefore, we have excluded cases below 45 years and transgender cases. Hence, our study has taken 1051, 1628 and 1814 deceased individual cases from the 60^th^, 71^st^ and 75^th^ rounds respectively. Thus, the pooled sample consists of 4493 deceased adults in the study.

### Outcome variable

The outcome variable in the study is a dichotomous. This is for the particular former Household member who died in the last 365 days prior to the survey. In the questionnaire (Block 5, NSSO), it has been asked “whether medical attention received before death?” Yes=1 and NO=2. Therefore, our study has categorized the outcome variable as:

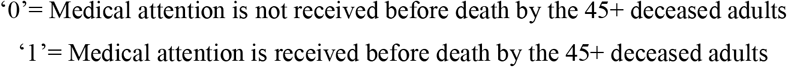

### Independent variables

Gender is classified as male and femlae while age-groups is categorized into three such as middle-aged adults which is 45-59 years, young-old which is 60-69 years age groups, middle-old is 70-79 years, oldest-old is 80+ years. Residence is categorized into two such as rural and urban. Regions is classified into six geographical zones such as Northern (includes Jammu and Kashmir, Delhi, Haryana, Himachal Pradesh, Chandigarh, Punjab, Uttarakhand, and Rajasthan), North-eastern (includes Assam, Arunachal Pradesh, Meghalaya, Manipur, Nagaland, Mizoram, Tripura and Sikkim), Central (includes Madhya Pradesh, Uttar Pradesh, and Chhattisgarh), Eastern (includes Odisha, Bihar, Jharkhand, West Bengal), Western (includes Goa, Maharashtra, Gujarat, Daman & Diu, and Dadra & Nagar Haveli), and Southern (includes Tamil Nadu, Kerala, Telangana, Andhra Pradesh, Karnataka, Puducherry, Andaman and Nicobar Islands, and Lakshadweep). In case of religion, it has two main categories such as Non-Hindus (includes Christians, Buddhist, Sikhs, Jews, Parsis etc.), and Hindus. We have also used caste groups which has three groups such as Schedule caste (SC) & Schedule Tribe (ST), Other backward caste (OBC) and general (no caste). Household’s wealth index which is categorized into five, such as Poorest, Poorer, Middle, Richer and Richest respectively. The household size is consists of two, such as-less than or equal to five-members and more than five members. Our study has also adopted household’s working type such as casual-workers, regular-wages, self-employed and others respectively.

### Analysis

Our study has performed univariate, bivariate and multivariate analyses. First, we have obtain the descriptive results where the sample distribution of the 45+ deceased adults with suitable background characteristics are calculated. Second, we have estimated the trends in meidcal attention rate received before death by the 45+ deceased adults with suitable background characteristics for the time period 2004-2005, 2014 and 2017-2018 respectively. Third, we have estimated the absolute changes in medical attention rate (MAR) received before death by the 45+ deceased adults for the time period between 2004-2005 & 2017-2018 and 2014 & 2017-2018. These canges can be written as:

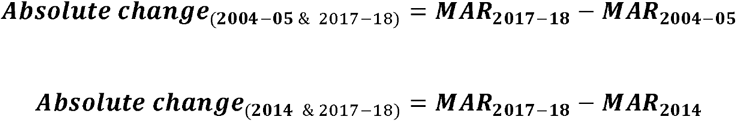

Lastly, we have performed binary logistic regression analysis to identify the effect of significant predictor variables on the outcome variable i.e., medical attention received before death among 45+ deceased adults. The regression model can be defined as in the following equation:

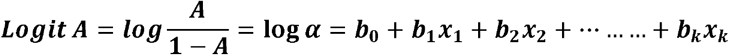

Where, ‘A’ is the probability (event Y occurs); ‘b’ is the factor (odds changes with unit increase in the independent variable); If ‘b’>0, then the odds ratio (OR) will increase and the factor ‘b’ >1. On the other hand, if ‘b’<0, then the odds ratio will decrease. And if ‘b’=0, then the odds remain unchanged.

## Results

### Sample distribution of 45+ deceased adults in India

Table 1 presents the sample distribution of the 45+ deceased adults in India using pooled-cross-sectional survey data from 2004 to 2018. The percentage share of 45+ deceased females (37.57%) is found to lower compared to 45+ males (62.43%). Middle-aged adults (28.98%) have the highest percentage share of deceased adults followed by Young-old (28.15%) and Middle-old (24.33%) while Oldest-old (18.54%) has the lowest. The percentage share of 45+ deceased adults are greater in Rural residence (55.95%) than Urban (44.05%). Southern regions (20.7%) showing the highest percentage share of 45+ deceased adults followed by Central (20.3%), Eastern (19.88%) and Northern (17.03%) while North-eastern (10.02%) has the lowest. Around 78.37% of 45+ deceased adults belong to Hindu religion. OBC caste (37.81%) has highest percentage share of 45+ deceased adults, while General caste (36.43%) has slightly lower than OBC but SC/ST has the lowest 25.63%. Interestingly, the percentage share of 45+ deceased adults is found to be greatest among Richest wealth group with 30.22% and Poorer has the lowest with only 15.13%. The household size with more than five members (53.05%) and Self-employed household working type are indicating highest 45+ deceased adults.

**Table 1.** Sample distribution of the 45+ deceased individuals in India using three pooled cross-sectional survey data from 2004 to 2018.

| Background characteristics | 2004-2005<br>(A) |  | 2014<br>(B) |  | 2017-2018<br>(C) |  | Pooled<br>(A+B+C=N) |  |
| --- | --- | --- | --- | --- | --- | --- | --- | --- |
|  | Percentage (%) | Numbers | Percentage (%) | Numbers | Percentage (%) | Numbers | Percentage (%) | Numbers |
| **Gender** |  |  |  |  |  |  |  |  |
| Male | 58.8 | 618 | 63.33 | 1,031 | 63.73 | 1,156 | 62.43 | 2,805 |
| Female | 41.2 | 433 | 36.67 | 597 | 36.27 | 658 | 37.57 | 1,688 |
| **Age-groups** |  |  |  |  |  |  |  |  |
| Middle-aged |  |  |  |  |  |  |  |  |
| adult | 28.45 | 299 | 29.24 | 476 | 29.05 | 527 | 28.98 | 1,302 |
| Young-old | 24.45 | 257 | 28.07 | 457 | 30.37 | 551 | 28.15 | 1,265 |
| Middle-old | 25.4 | 267 | 24.82 | 404 | 23.26 | 422 | 24.33 | 1,093 |
| Oldest-old | 21.69 | 228 | 17.87 | 291 | 17.31 | 314 | 18.54 | 833 |
| **Residence** |  |  |  |  |  |  |  |  |
| Rural | 68.7 | 722 | 51.04 | 831 | 52.98 | 961 | 55.95 | 2,514 |
| Urban | 31.3 | 329 | 48.96 | 797 | 47.02 | 853 | 44.05 | 1,979 |
| **Regions** |  |  |  |  |  |  |  |  |
| Northern | 14.27 | 150 | 14.37 | 234 | 21 | 381 | 17.03 | 765 |
| North-eastern | 12.46 | 131 | 10.57 | 172 | 8.1 | 147 | 10.02 | 450 |
| Central | 20.17 | 212 | 19.72 | 321 | 20.89 | 379 | 20.3 | 912 |
| Eastern | 19.98 | 210 | 19.04 | 310 | 20.56 | 373 | 19.88 | 893 |
| Western | 10.94 | 115 | 12.78 | 208 | 12.13 | 220 | 12.09 | 543 |
| Southern | 22.17 | 233 | 23.53 | 383 | 17.31 | 314 | 20.7 | 930 |
| **Religion** |  |  |  |  |  |  |  |  |
| Non-Hindu | 20.84 | 219 | 20.64 | 336 | 22.99 | 417 | 21.63 | 972 |
| Hindu | 79.16 | 832 | 79.36 | 1,292 | 77.01 | 1,397 | 78.37 | 3,521 |
| **Caste** |  |  |  |  |  |  |  |  |
| SC/ST | 26.74 | 281 | 25.31 | 412 | 25.58 | 464 | 25.75 | 1,157 |
| OBC | 38.63 | 406 | 40.05 | 652 | 35.34 | 641 | 37.81 | 1,699 |
| General | 34.63 | 364 | 34.64 | 564 | 39.08 | 709 | 36.43 | 1,637 |
| Wealth Index |  |  |  |  |  |  |  |  |
| Poorest | 15.89 | 167 | 14.99 | 244 | 16.43 | 298 | 15.78 | 709 |
| Poorer | 13.61 | 143 | 18 | 293 | 13.45 | 244 | 15.13 | 680 |
| Middle | 19.51 | 205 | 16.46 | 268 | 15.88 | 288 | 16.94 | 761 |
| Richer | 21.79 | 229 | 18.8 | 306 | 24.81 | 450 | 21.92 | 985 |
| Richest | 29.21 | 307 | 31.76 | 517 | 29.44 | 534 | 30.22 | 1,358 |
| **HH Size** |  |  |  |  |  |  |  |  |
| <=5 | 38.73 | 407 | 47.17 | 768 | 51.49 | 934 | 46.94 | 2,109 |
| >5 | 61.27 | 644 | 52.83 | 860 | 48.51 | 880 | 53.06 | 2,384 |
| **HH Type** |  |  |  |  |  |  |  |  |
| Casual workers | 23.5 | 247 | 17.44 | 284 | 13.78 | 250 | 17.38 | 781 |
| Regular-wages | 9.71 | 102 | 24.08 | 392 | 11.58 | 210 | 15.67 | 704 |
| Self-employed | 53.76 | 565 | 49.69 | 809 | 64.88 | 1,177 | 56.78 | 2,551 |
| Others | 13.04 | 137 | 8.78 | 143 | 9.76 | 177 | 10.17 | 457 |
| **Total** | **100** | **1,051** | **100** | **1,628** | **100** | **1,814** | **100** | **4,493** |

Source: Author's own calculation
Abbreviations: SC-Schedule caste; ST-Schedule tribe; OBC-Other backward caste

### Trends in medical attention rate (MAR) by 45+ deceased adults in India

Figure 1 presents the trend in medical attention rate received before death by 45+ deceased adults from 2004 to 2018 in India. The result shows an increasing trend in MAR among males from 2004-05 to 2014 and then it is dropped by 5.4% in 2017-18 while females have showed declining trend from 2004-2018. Middle-aged & Oldest-old adults have showed increasing trends in MAR while Young-old & Middle-old are indicating decreasing MAR trend from 2004-2018. The MAR in rural residence has been dropped to 2.21 % whereas the MAR for urban residence has been rose to 6.09% from 2004 to 2017-18. The MAR trend is declined in North-eastern by 9.35%, Central by 3.04% & Southern regions by 2.44% while Northern by 11.61%, Eastern by 5.59% & Western by 1.32% indicating increasing trend but spike is reflected by Western regions in 2014. Trend in MAR among Hindu religion is increased by 2.03% whereas trends in MAR among Non-Hindu is dropped by 4.6%. General caste is showing increasing trend in MAR while both SC/ST & OBC are showing declining trend in MA. However, the trends in MAR among Poorest is dropped by 14.14% while both Middle & Richer are showing marginally increasing trend but the trends in MAR is increased among both Poorer by 6.56% & Richer by 12.13%. The trend in MAR among HH-member less than five is increased by 2.1%. Casual-workers and regular-wages are showing declining trends in MAR while Self-employed & Others are indicating increasing trends.

**Figure 1.**
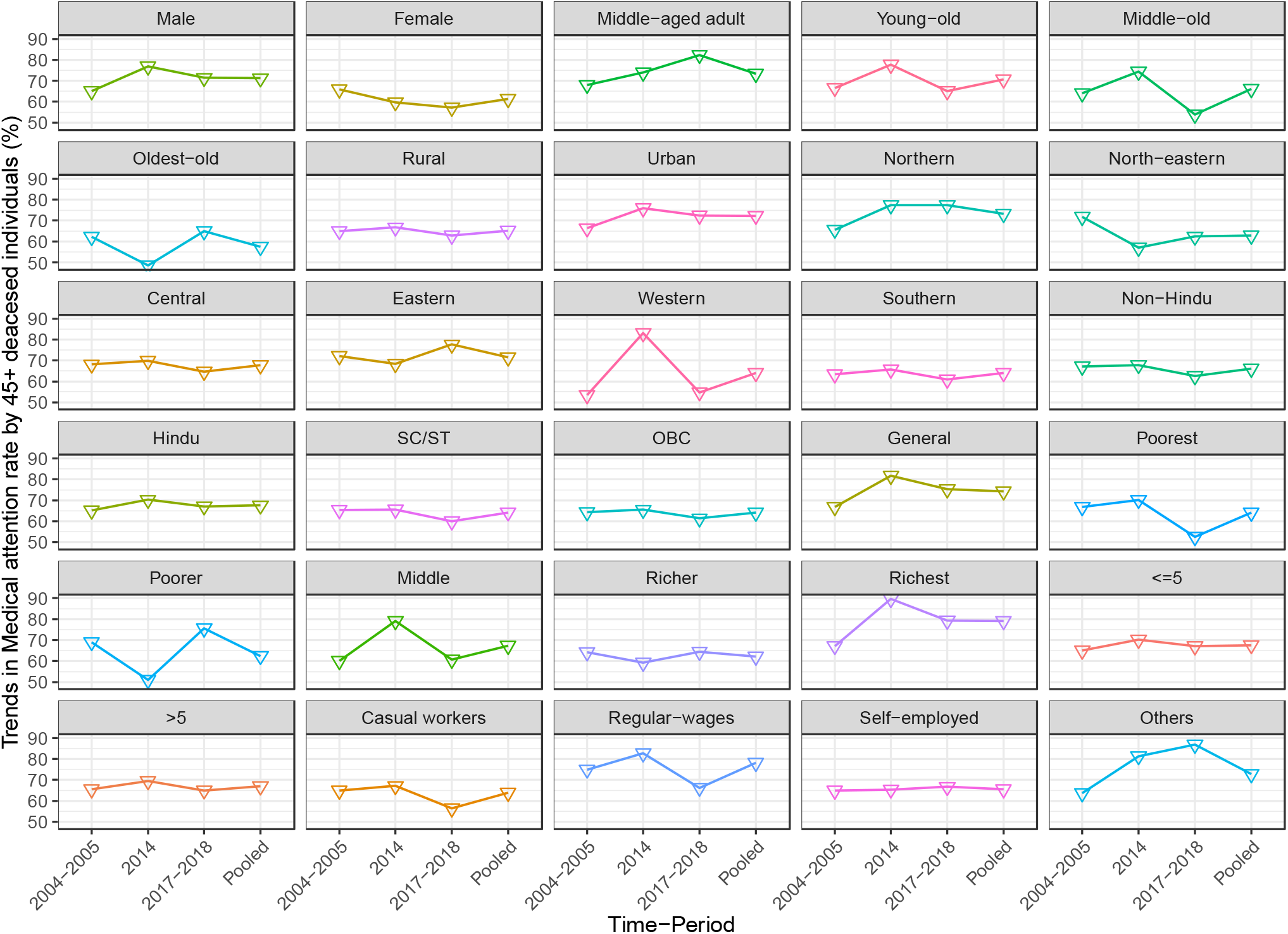
Trends in medical attention rates (%) received before death by 45+ individuals in India with the suitable background characteristics, from 2004 to 2018.

### Changes in medical attention rate (MAR) by 45+ deceased adults in India between 2004-2018 and 2014-2018

Figure 2 presents the changes in medical attention rate (MAR) received by 45+ deceased adults in India between 2004 & 2018 and 2014-2018. The overall absolute change in MAR between 2004-2018 is positive with an increment of 0.74%. The absolute positive change in MAR (2004-2018) is seen among males with 6.39%, while females show absolute negative change with −8.67%. Middle-aged adults (14.29%) and oldest-old (2.57%) are showing absolute positive changes in MAR between 2004 and 2018, while young-old (−1.5%) and middle-old (−10.14%) are indicating negative changes. Rural residence (−2.21%) has negative changes in MAR (2004-2018), whereas positive change is reflected in urban residence (6.09%) respectively. In the case of regions, Northern (11.61%), Eastern (5.59%), and Western (1.32%) are showing absolute positive changes in MAR (2004-2018) while the North-eastern (−9.35%), Central (−3.47%) and Southern (−2.44%) are reflecting negatively. Non-Hindus (−4.6%) religion indicates an absolute negative change in MAR between 2004 and 2018, whereas Hindus have positive change with 2.03%. MAR has increased among General (8.48%) caste compared to SC/ST and OBC between 2004 and 2018. Except poorest, all other wealth groups are showing positive changes, which means the MAR has increased between 2004-2018. Household size with more than five members (−0.6%) showing a negative change in MAR while <= five members (2.1%) is indicating positive.

**Figure 2.**
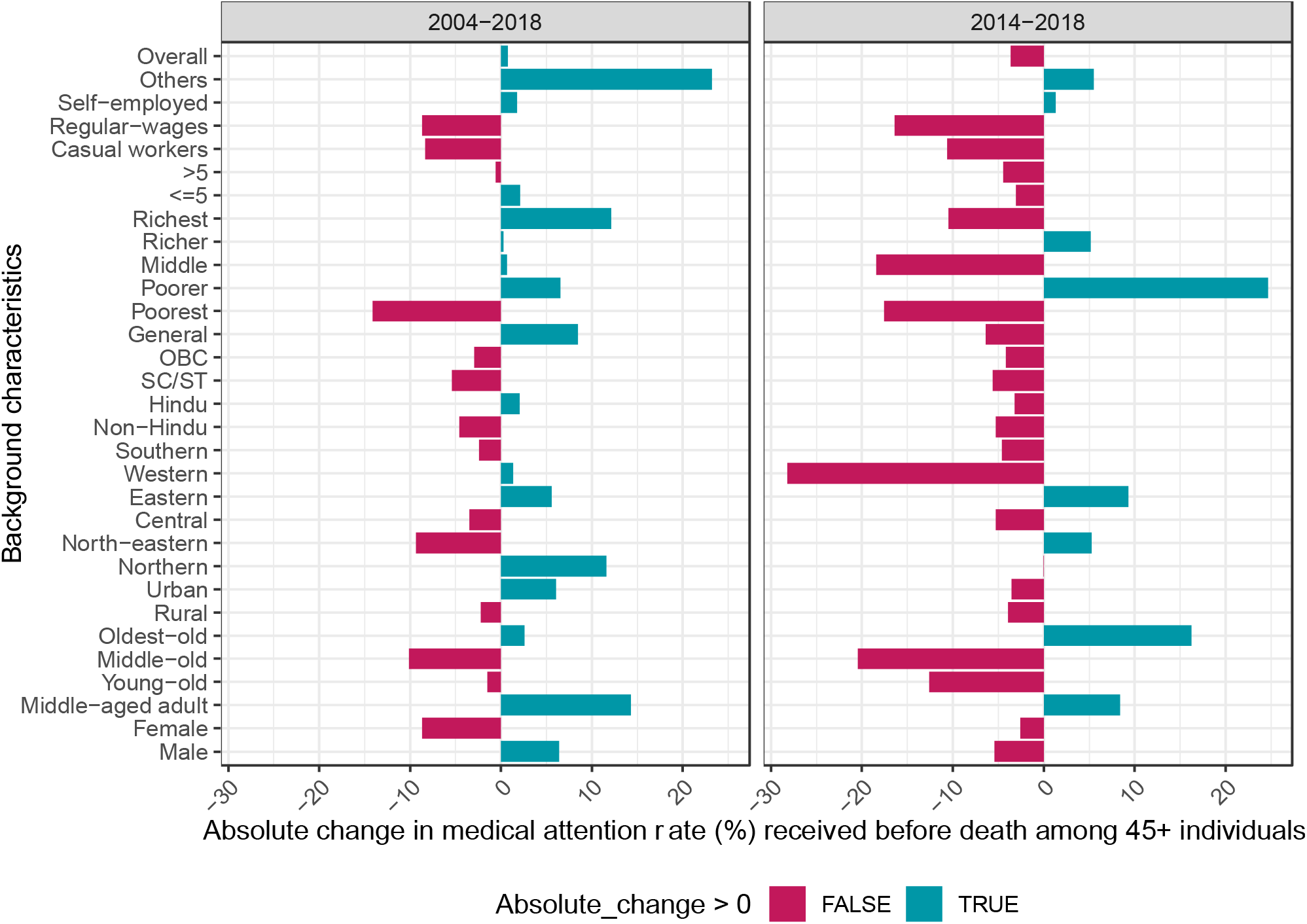
Absolute change (%) in medical attention received before death by 45+ individuals in India with suitable background characteristics for the time period between 2004 & 2018 and 2014 & 2018. Source: Author’s own calculation Abbreviations: SC-Schedule caste; ST-Schedule tribe; OBC-Other backward caste

However, a negative overall absolute change in MAR is observed with a decrement of −3.68% between 2014-2018. The result shows that both males (−5.44%) and females (−2.57%) show an absolute negative change in MAR between 2014-2018. Middle-aged (8.41%) adults and oldest-old (16.24%) are indicating a positive change in MAR between 2014-2018 while young-old (−12.62%) and middle-old (−20.46%) are showing negative, thus similar absolute changes are observed between 2004-2018. Both rural and urban residences are observed to have negative changes, which means the MAR is declined by −3.91% in rural and −3.53% in urban residences. Interestingly, the absolute change in MAR is negative for Northern (− 0.02%), Central (−5.27%), Western (−28.22%) and Southern (−4.63%) regions, whereas North-eastern (5.26%) and Eastern (9.31%) regions are showing positive changes. Hindus, non-Hindus, SC/ST, OBC and General are showing negative changes in MAR between 2014- 2018. Poorer and richer wealth groups have a positive change, which means their MAR has increased between 2014-2018, whereas the poorest, middle and richest have negative changes. Both households with <= five members and >5members show negative changes in MAR. Though, self-employed (1.3%) and others (5.48%) household working type are showing positive changes in MAR between 2014-2018 while casual-workers (−10.62%) and regular-wages (−16.43%) are indicating negative changes respectively.

### Regression results of medical attention received before death by 45+ adults in India

Table 2 presents the binary logistics regression results of medical attention received before death by 45+ deceased adults in India using pooled-cross-sectional survey data (2004-2018). Young-old (Odds=0.82; p<0.1), middle-old (Odds=0.6; p<0.01) and oldest-old (Odds=0.43; p<0.01) are showing significantly lower odds of receiving medical attention before death compared to middle-aged adults. Surprisingly, Southern (Odds=0.72; p<0.05) and western (Odds=0.68; p<0.01) regions are found to have significantly lower odds of receiving medical attention before death but Eastern (Odds=1.75; p<0.01) region has higher odds in compared to Northern region. Greater odds of receiving medical attention before death is reflected among General caste (Odds=1.26; p<0.05) than SC/ST. Both richer (Odds=1.51; p<0.01) and richest (Odds=1.71; p<0.01) wealth groups are showing significantly higher odds of receiving medical attention before death compared to Poorest. The survey year 2014 (Odds=3.39; p<0.01) and survey year 2017-2018 (Odds=2.07; p<0.01) are found to have significantly greater odds of receiving medical attention before death compared to the survey year 2004-2005.

**Table 2.** The logistic regression result showing the medical attention received before death by 45+ individual in India using pooled-cross-sectional survey data from 2004-2018 (Pooled sample=4493).

| Background characteristics |  | Odds ratio<br>(OR) | Lower Conf.<br>Interval<br>(LC) | Upper Conf.<br>Interval<br>(UC) | p-value |
| --- | --- | --- | --- | --- | --- |
| **Gender** | Male | 1 |  |  |  |
|  | Female | 1.04 | 0.88 | 1.22 | 0.664 |
| **Age-groups** | Middle-aged adult | 1 |  |  |  |
|  | Young-old | 0.82* | 0.65 | 1.03 | 0.086 |
|  | Middle-old | 0.60*** | 0.48 | 0.75 | 0 |
|  | Oldest-old | 0.43*** | 0.34 | 0.55 | 0 |
| **Residence** | Rural | 1 |  |  |  |
|  | Urban | 1.16 | 0.96 | 1.40 | 0.121 |
| **Regions** | Northern | 1 |  |  |  |
|  | North-eastern | 0.90 | 0.65 | 1.23 | 0.502 |
|  | Central | 1.13 | 0.85 | 1.49 | 0.398 |
|  | Eastern | 1.75*** | 1.30 | 2.36 | 0 |
|  | Western | 0.68*** | 0.51 | 0.91 | 0.01 |
|  | Southern | 0.72** | 0.55 | 0.94 | 0.016 |
| **Religion** | Non-Hindu | 1 |  |  |  |
|  | Hindu | 1.14 | 0.94 | 1.38 | 0.192 |
| **Caste** | SC/ST | 1 |  |  |  |
|  | OBC | 1.02 | 0.83 | 1.26 | 0.83 |
|  | General | 1.26** | 1.00 | 1.57 | 0.045 |
| **Wealth index** | Poorest | 1 |  |  |  |
|  | Poorer | 1.03 | 0.78 | 1.35 | 0.859 |
|  | Middle | 1.13 | 0.86 | 1.48 | 0.386 |
|  | Richer | 1.51*** | 1.14 | 1.98 | 0.004 |
|  | Richest | 1.71*** | 1.28 | 2.30 | 0 |
| **HH members** | <=5 | 1 |  |  |  |
|  | >5 | 1.02 | 0.86 | 1.22 | 0.792 |
| **HH working type** | Casual workers | 1 |  |  |  |
|  | Regular-wages | 1.04 | 0.78 | 1.40 | 0.774 |
|  | Self-employed | 1.06 | 0.85 | 1.32 | 0.61 |
|  | Others | 1.14 | 0.82 | 1.59 | 0.422 |
| **Year** | 2004-2005 | 1 |  |  |  |
|  | 2014 | 3.39*** | 2.74 | 4.19 | 0 |
|  | 2017-2018 | 2.07*** | 1.71 | 2.51 | 0 |

Source: Author's own calculation
Notes: Significant levels \*\*\*p-value<0.01; \*\*p-value<0.05; \*p-value<0.1
Abbreviations: SC-Schedule caste; ST-Schedule tribe; OBC-Other backward caste; LC-Lower confidence interval; UC-Upper confidence interval.

## Discussion

Our study has revealed that there has been marginal increase (0.74%) in the overall trend in medical attention received before death among 45+ adults from 2004 to 2017-18. On the other hand, the recent SRS Report (2019) has documented an increase in the medical attention for all ages in India; to be precise from 42.6% in 2014 to 47.8% in 2018. Though, our research focuses on a critical issue of 45+ adults in India that has yet to receive the attention it deserves. As a result, much study and debate focus on morbidity and mortality, with little mention of the reality that a huge majority of deaths still occur without medical intervention, despite the fact that death statistics do not reflect this.

Interestingly, our finding revealed that females are showing negative change in receiving medical attention than male and the odds of receiving medical attention is higher among female than males but did not show significant while similar results have been also reflected in other studies (Ghosh & Sengupta, 2016; Gupta & Sankar, 2004). Though, female lifespan has much improved in India compared but still the heterogeneity exists in seeking health care as they are more economically dependent on others and financially vulnerable too (Akhtar & Saikia, 2022; Maharana & Ladusingh, 2014; Moradhvaj & Saikia, 2019; Saikia, Moradhvaj, et al., 2016). Furthermore, there are not many community-based health centers that is dedicated solely to women’s needs. Additionally, there is a dearth of an age-appropriate healthcare support system exacerbates the challenges. Previous studies suggest that the disparities exist in access to health care among middle-aged and older adults (Mair et al., 2016; McGrath et al., 2019; Miller et al., 2014) and our finding has showed that young-old, middle-old and oldest-old have significantly lower odds of receiving medical attention compared to middle-aged adults.

Nevertheless, in general, there is a vast difference between rural and urban residence in terms of treatment seeking behavior, our finding has revealed that medical attention rate is lower in rural residence compared to urban and the previous study also evident the similar results (Ghosh & Sengupta, 2016; Gupta & Sankar, 2004). Although the public health care institutes is improving now in rural areas but still it lacks the quality of advance medical facilities in the rural India which also well documented (Bose, 2005). However, in India, a better public health services exists, but it continues to be hampered by issues of accessibility, availability, and quality, particularly in rural areas. When these are paired with issues of poverty, underdevelopment and lack of knowledge and awareness, it leads to a lack of medical care prior to death.

Surprisingly, our result has revealed that Southern and Western regions are found to have significantly less likely to receive medical attention before death among deceased adults which is a striking finding. While the previous studies have found that older adults belong to Southern region were more likely receive hospitalizations (Akhtar & Saikia, 2022; Banerjee, 2021; Giridhar et al., 2014; Pandey et al., 2017a, 2017b). In contrast, the recent SRS report (2019) found that, with the exception of Kerala, all other southern states (Tamil Nadu, Telangana, Karnataka, and Andhra Pradesh) had below the national average percentage of medical attention before death (SRS Report, 2019). On the other hand, our finding has also revealed that Eastern region is significantly more likely to receive medical attention before death which is also an exceptional result while previous studies showed a contradictory results (Akhtar & Saikia, 2022; Pandey et al., 2017a, 2017b). Even, the SRS report (2019) has documented that except Odisha, all other easter states (West Bengal, Bihar and Jharkhand) had lower percentage of receiving medical attention before death for all ages than the national average (SRS Report, 2019).

Meanwhile, our finding also identified that richer and richest household income group is significantly higher impact in receiving medical attention compared to poorest and it is also reflected in the earlier researches (Ghosh & Sengupta, 2016; Gupta & Sankar, 2004). As a result, it is apparent that the poorest segment suffers the most in terms of receiving medical care. Our finding revealed that those deceased adults belonging to General caste category are significantly more likely to receive medical attention before death compared to SC/ST caste and the previous results also found the similar results (Akhtar & Saikia, 2022; Ghosh & Sengupta, 2016; Gupta & Sankar, 2004). However, our finding also revealed that religion did not play a key role in determining the who gets medical attention before death.

## Limitations and Strengths

Our study has several limitations. First, we have used a nationally pooled-cross-sectional survey data which cannot establish causality or rule out reciprocal causation in our findings, Secondly, other significant factors can also predict medical attention and provide insightful results, such as health behavioral factors, health beliefs, perceptions, lifestyle factors, obesity, mental state, educational level, marital status, living arrangements, insurance policy coverage and several personal habits like smoking cigarettes, drinking alcohol, consuming tobacco or other harmful substances. However, these information’s are not available in the survey data, therefore we could not include it.

The strengths of the present study include the nationally representative pooled-cross-sectional survey data from 2004 to 2017-18 which allowed us to quantify trends, changes and factors associated with the medical attention. It is the first study which broadly focused on 45+ deceased adults (middle-aged and older adults). Our study has considered a broad range of factors, including socio-demographic and economics characteristics of the deceased adults. Future research should look into other aspects that may be linked to avoidance of medical attention in an older population, such as the impact of preconceptions about aging and old age on care seeking, the impact of multi-morbidities conditions and symptoms perception on care seeking, or the impact of future temporal perspectives.

## Conclusions

Therefore, there is an urgent need to establish a primary health center at community level that play an important role in order to meet the comprehensive requirements of middle-aged and older adults in India. It would be helpful to develop and execute the health policies and programs that would enable India to achieve healthy aging in India at national and subnational levels, as it is a key element of public health challenges. The hospitals and other health-care institutes should ensure that public health measures and actions for middle-aged and older adults must aligned with those in health care and community-level services sectors at local, districts, and state levels.

## Data Availability

The data is publicly available and can be accessed at https://www.mospi.gov.in/web/mospi/download-tables-data/-/reports/view/templateTwo/16202?q=TBDCAT

## Acknowledgements

None

## References

Akhtar, S. N., & Saikia, N. (2022). Differentials and predictors of hospitalisation among the elderly people in India: Evidence from 75th round of National Sample Survey (2017-2018). Working with Older People, ahead-of-print(ahead-of-print). https://doi.org/10.1108/WWOP-11-2021-0055

Alam, N., Chowdhury, H. R., Bhuiyan, M. A., & Streatfield, P. K. (2010). Causes of Death of Adults and Elderly and Healthcareseeking before Death in Rural Bangladesh. Journal of Health, Population and Nutrition, 28(5), 520–528. https://doi.org/10.3329/jhpn.v28i5.6161

Ali, I., Akhtar, S. N., Chauhan, B. G., Malik, M. A., & Singh, K. D. (2022). Health insurance support on maternal health care: Evidence from survey data in India. Journal of Public Health, fdac025. https://doi.org/10.1093/pubmed/fdac025

Banerjee, S. (2021). Determinants of rural-urban differential in healthcare utilization among the elderly population in India. BMC Public Health, 21(1), 939. https://doi.org/10.1186/s12889-021-10773-1

Bhat, M. (1987). MORTALITY IN INDIA: LEVELS, TRENDS AND PATTERNS [University of Pennsylvania]. https://repository.upenn.edu/dissertations/AAI8725140

Bora, J. K., & Saikia, N. (2018). Neonatal and under-five mortality rate in Indian districts with reference to Sustainable Development Goal 3: An analysis of the National Family Health Survey of India (NFHS), 2015–2016. PLOS ONE, 13(7), e0201125. https://doi.org/10.1371/journal.pone.0201125

Boro, B., Srivastava, S., & Saikia, N. (2021). Is There an Association Between Change in Old-Age Living Arrangements and Older Adults’ Psychological Health and Subjective Well-Being in India? Evidence from a Cross-Sectional Study. Ageing International. https://doi.org/10.1007/s12126-021-09470-6

Bose, A. (2005). Private health sector in India: Is private health care at the cost of public health care? BMJ (Clinical Research Ed.), 331(7528), 1338–1339. https://doi.org/10.1136/bmj.331.7528.1338-c

Chauhan, S., Srivastava, S., Kumar, P., & Patel, R. (2022). Decomposing urban-rural differences in multimorbidity among older adults in India: A study based on LASI data. BMC Public Health, 22(1), 502. https://doi.org/10.1186/s12889-022-12878-7

Gbeasor-Komlanvi, F. A., Tchankoni, M. K., Bakoubayi, A. W., Lokossou, M. Y., Sadio, A., Zida-Compaore, W. I. C., Djibril, M., Belo, M., Agbonon, A., & Ekouevi, D. K. (2020). Predictors of three-month mortality among hospitalized older adults in Togo. BMC Geriatrics, 20(1), 507. https://doi.org/10.1186/s12877-020-01907-y

Ghosh, D., & Sengupta, J. (2016). Medical Intervention before Death of the Elderly in India. SSRN Electronic Journal. https://doi.org/10.2139/ssrn.2811049

Giridhar, G., Sathyanarayana, K. M., Kumar, S., James, K. S., & Alam, M. (Eds.). (2014). Population Ageing in India. Cambridge University Press. https://doi.org/10.1017/CBO9781139683456

Gupta, I., & Sankar, D. (2004). Medical Attention at Death: Evidence from India. Journal of Health Management, 6(1), 73–84. https://doi.org/10.1177/097206340400600105

Kannan, V. D., & Veazie, P. J. (2014). Predictors of Avoiding Medical Care and Reasons for Avoidance Behavior. Medical Care, 52(4), 336–345. https://doi.org/10.1097/MLR.0000000000000100

Khan, M. R., Malik, M. A., Akhtar, S. N., & Yadav, S. (2021). Multimorbidity and its associated risk factors among the older adults in India [Preprint]. Public and Global Health. https://doi.org/10.1101/2021.11.12.21265083

Kumar, S., & Kumar, K. A. (2022). Socio-economic and Demographic Correlates of the Health Status of Older Adults in India: An Analysis of NSS 71st Round Data. Ageing International, 47(1), 89–112. https://doi.org/10.1007/s12126-020-09407-5

Kundu, J., Mishra, P. S., & Bharadwaz, M. P. (2022). Predictors of Healthcare Utilization among Geriatrics in India: Evidence from National Sample Survey Organization, 2017–18. Ageing International. https://doi.org/10.1007/s12126-021-09481-3

Law Insider. (2022). Definition of Medical attention. https://www.lawinsider.com/dictionary/medical-attention

Leyva, B., Taber, J. M., & Trivedi, A. N. (2020). Medical Care Avoidance Among Older Adults. Journal of Applied Gerontology, 39(1), 74–85. https://doi.org/10.1177/0733464817747415

Maharana, B., & Ladusingh, L. (2014). Gender Disparity in Health and Food Expenditure in India among Elderly. International Journal of Population Research, 2014, 1–8. https://doi.org/10.1155/2014/150105

Mair, C. A., Quiñones, A. R., & Pasha, M. A. (2016). Care Preferences Among Middle-Aged and Older Adults With Chronic Disease in Europe: Individual Health Care Needs and National Health Care Infrastructure. The Gerontologist, 56(4), 687–701. https://doi.org/10.1093/geront/gnu119

McGrath, R., Al Snih, S., Markides, K., Hall, O., & Peterson, M. (2019). The burden of health conditions for middle-aged and older adults in the United States: Disability-adjusted life years. BMC Geriatrics, 19(1), 100. https://doi.org/10.1186/s12877-019-1110-6

Miller, N. A., Kirk, A., Kaiser, M. J., & Glos, L. (2014). Disparities in Access to Health Care Among Middle-Aged and Older Adults with Disabilities. Journal of Aging & Social Policy, 26(4), 324–346. https://doi.org/10.1080/08959420.2014.939851

Moradhvaj, & Saikia, N. (2019). Gender disparities in health care expenditures and financing strategies (HCFS) for inpatient care in India. SSM - Population Health, 9, 100372. https://doi.org/10.1016/j.ssmph.2019.100372

Pandey, A., Ploubidis, G. B., Clarke, L., & Dandona, L. (2017a). Horizontal inequity in outpatient care use and untreated morbidity: Evidence from nationwide surveys in India between 1995 and 2014. Health Policy and Planning, 32(7), 969–979. https://doi.org/10.1093/heapol/czx016

Pandey, A., Ploubidis, G. B., Clarke, L., & Dandona, L. (2017b). Hospitalisation trends in India from serial cross-sectional nationwide surveys: 1995 to 2014. BMJ Open, 7(12), e014188. https://doi.org/10.1136/bmjopen-2016-014188

Prinja, S., Kaur, M., & Kumar, R. (2012). Universal health insurance in India: Ensuring equity, efficiency, and quality. Indian Journal of Community Medicine: Official Publication of Indian Association of Preventive & Social Medicine, 37(3), 142–149. https://doi.org/10.4103/0970-0218.99907

Sahoo, H., Govil, D., James, K. S., & Prasad, R. D. (2021). Health issues, health care utilization and health care expenditure among elderly in India: Thematic review of literature. Aging and Health Research, 1(2), 100012. https://doi.org/10.1016/j.ahr.2021.100012

Saikia, N. Moradhvaj, & Bora, J. K. (2016). Gender Difference in Health-Care Expenditure: Evidence from India Human Development Survey. PLOS ONE, 11(7), e0158332. https://doi.org/10.1371/journal.pone.0158332

Saikia, N., Shkolnikov, V. M., Jasilionis, D., & Chandrashekhar. (2016). Trends and Sub-National Disparities in Neonatal Mortality in India from 1981 to 2011. Asian Population Studies, 12(1), 88–107. https://doi.org/10.1080/17441730.2015.1130325

Spleen, A. M., Lengerich, E. J., Camacho, F. T., & Vanderpool, R. C. (2014). Health Care Avoidance Among Rural Populations: Results From a Nationally Representative Survey: Health Care Avoidance Among Rural Populations. The Journal of Rural Health, 30(1), 79–88. https://doi.org/10.1111/jrh.12032

SRS Report. (2019). Sample registration System Report. Office of Registrar general, Census Commissioner, India, Ministry of Home Affairs. https://censusindia.gov.in/Vital_Statistics/SRS_Report_2019/SRS%20Statistical%20Report%202019.pdf

